# Current international tools and guidance for the implementation of hand hygiene recommendations in community settings: a scoping review

**DOI:** 10.1101/2024.05.24.24307876

**Authors:** Clara MacLeod, Joanna Esteves Mills, Bethany A Caruso, Claire Chase, Kondwani Chidziwisano, Jenala Chipungu, Robert Dreibelbis, Regina Ejemot-Nwadiaro, Bruce Gordon, Ann Thomas, Oliver Cumming, Laura Braun

## Abstract

**Background:** Hand hygiene is an important measure to prevent disease transmission. This scoping review identifies and summarises current tools and guidance for the implementation of hand hygiene recommendations in community settings.

**Methods:** We conducted a scoping review following the Arksey and O’Malley framework. To identify relevant documents, we searched: 1) a grey literature database, 2) Google search engine, and 3) the websites of international organisations in August 2023. We included tools and implementation guidance relevant to hand hygiene in community settings, categorised as domestic, public, or institutional, and published in English by international organisations between January 1990 and August 2023. Tools and implementation guidance were mapped to an existing conceptual framework adapted for this review that includes a six-step implementation approach.

**Results:** We included a total of 35 documents, comprising 30 implementation guidance documents and 5 stand-alone tools. Among these 35 documents, we identified 207 implementation recommendations and a total of 21 tools for the six implementation steps. The 21 tools include 5 stand-alone tools and 16 tools embedded within guidance documents. Most implementation guidance was mapped to steps 1 (prepare for action), 2 (analyse the situation), 3 (develop an action plan), and 5 (monitor, evaluate, and course correct) of the conceptual framework, with limited guidance for step 4 (executing the action plan) and step 6 (cross-cutting themes). Over half of identified tools are for step 2 (analyse the situation) and primarily for undertaking a situation analysis. Only two documents provided guidance or a tool across the six steps.

**Conclusion:** Implementation guidance is available, yet inconsistently spread across the different implementation steps. There is also a limited number of tools to support implementation. Future work should focus on developing comprehensive practical tools for the implementation of hand hygiene recommendations in community settings, in line with international guidelines.

## Introduction

Hand hygiene, which includes handwashing with soap and the use of alcohol-based hand rub (ABHR), is an important public health measure for the control and prevention of infectious diseases (1–3). Handwashing with soap has been found to be a cost-effective intervention that can reduce the risk of both diarrhoeal disease and acute respiratory infections by over 20% (4–7). Despite the international recognition of hand hygiene as a critical public health measure, there continues to be insufficient access to products and basic services in community settings, particularly in low- and middle-income countries (8–10). A recent global hygiene assessment found that most surveyed countries had national policies for hand hygiene, but over a third did not have a financial plan for implementing them (11).

While clear and robust recommendations developed by the World Health Organization (WHO) exist for hand hygiene in health care settings, there are gaps in global normative guidance on hand hygiene in settings where health care is not routinely delivered (12). A recent scoping review of current international guidelines for hand hygiene in community settings – categorised as non-healthcare settings – highlighted a lack of consistent and evidence-based recommendations across four key areas: (1) what constitutes effective hand hygiene, (2) minimum requirements for practicing hand hygiene, (3) effective behaviour change approaches to sustain hand hygiene, and (4) the role of government (12). As part of its mandate to address demand for guidance on public health topics where there is uncertainty and demand from governments, WHO is developing Guidelines for hand hygiene in community settings, with a series of systematic reviews underway to inform this effort (13). The Guidelines will provide evidence-based recommendations on how to improve hand hygiene in non-healthcare settings, collectively referred to as community settings (including domestic, public and institutional settings) (14).

Implementation guidance and tools are critical for the uptake of the guidelines, as well as for translating recommendations into practice for guideline users. The WHO Guidelines will also include practical guidance on how to implement the guideline recommendations. This will be provided through step-by-step guidance and a set of accompanying tools to support adaptation of global recommendations into national action plans for hand hygiene in community settings (15). For the hand hygiene in health care guidelines, for example, WHO has published a guide to the implementation of the WHO multimodal hand hygiene improvement strategy. The guide is accompanied by several tools, including a planning and costing tool to help health-care facilities determine the feasibility of implementing alcohol-based handrub (16). Yet for the community setting, it is unclear what implementation guidance and tools currently exist. To our knowledge, this is the first review to summarise current tools and guidance for the global implementation of recommendations for hand hygiene in community settings. This review will provide a repository of tools and implementation guidance that can be used to address hand hygiene in community settings as part of WHO’s forthcoming Guidelines, as well as identify gaps for future work.

### Aim

The aim of this scoping review is to identify and summarise current implementation guidance and tools on hand hygiene in community settings to support the implementation of international recommendations.

## Methods

This review follows the six stages of the Arksey and O’Malley methodological framework for scoping reviews (17). Our review is described according to the Preferred Reporting Items for Systematic reviews and Meta-Analyses extension for Scoping Reviews (PRISMA-ScR) and a PRISMA-ScR checklist is included in the Supplementary Information Table S1. The protocol was preregistered with OSF Registries (18).

### Identifying the research question (stage 1)

Our primary research question is: “What current implementation guidance and tools are available to support the implementation of international recommendations for hand hygiene in community settings?”

#### Hand hygiene

For this review, hand hygiene refers to any hand cleansing undertaken for the purpose of removing or deactivating pathogens from hands and thereby limiting diseases transmission (19). The review considers implementation tools for planning, designing, executing, and monitoring and evaluating various aspects of hand hygiene interventions or programmes.

#### Implementation guidance and tools

We define an implementation guide as a document, such as a manual, handbook, or guide, that provides practical recommendations on how governments, non-governmental, and private sector actors can improve the uptake of hand hygiene in community settings in line with global guidelines (20). For this review, tools are defined as documents, such as checklists, worksheets, advocacy materials, costing spreadsheets, and education and training materials, that guideline users can use to practically implement guideline recommendations to improve hand hygiene in community settings (20). Tools are intended to provide users with a structured approach and decision-making support to ensure that recommended practices are effectively implemented. For example, there are several available tools related to planning, costing, and monitoring to guide users on the implementation of the guidelines on hand hygiene in health care (16,22,23). The WHO multimodal hand hygiene improvement strategy is another example of a tool to support the guideline implementation of hand hygiene in the healthcare setting (21). A tool may be included as part of a toolkit, here defined as a set of practical tools and resources to support hand hygiene improvement and sustainability, or a stand-alone tool. A stand-alone tool refers to a document that operates independently and is not integrated as part of a toolkit or broader document. Stand-alone tools provide targeted support for improving hand hygiene in community settings.

#### Community settings

This scoping review focuses on settings where health care is not routinely delivered (24), broadly spanning all places where people ‘learn, play, work and love’, referred to as ‘community settings’ (25). For this review, ‘community settings’ include: 1) domestic, 2) public, and 3) institutional settings globally (e.g., high-, medium-, and low-income countries). The domestic setting refers to households. The public setting includes markets, public transportation hubs, parks, squares, and other public spaces, such as shops and restaurants. The public setting also includes spaces that vulnerable populations, such as people experiencing homelessness, may occupy. Institutional settings include the workplace, schools and universities, places of worship, and prisons and places of detention (12).

#### Conceptual framework

To map the identified guidance and tools, we developed a conceptual framework, which includes a six-step approach to support the implementation of recommendations for hand hygiene in community settings (Figure 1). The conceptual framework also categorises community settings by domestic, public, and institutional setting. The six implementation steps (17,18) were adapted from the five-step approach from Burke et al. (2012) (26) and WHO (2009) (27) to include a cross-cutting sixth step. This approach has been utilised among WHO guidelines (28) based on the Centre for Effective Services (CES) Guide to Implementation (26). Step 1 (prepare for action) ensures the overall preparedness to design, implement and monitor a hand hygiene improvement plan. This includes appointing a lead ministry, identifying ministries responsible for each community setting of interest, establishing a coordination mechanism, and funding the development of a hand hygiene improvement plan. Step 2 (analyse the situation) aims to understand the national hand hygiene landscape for each community setting. This includes, where possible, information on hand hygiene practice, access to the minimum requirements, behaviour change programmes, and the enabling environment. Step 3 (develop an action plan) involves developing, costing, and financing a community setting-specific plan to implement a hand hygiene improvement programme based on findings from step 2. Step 4 (execute the action plan) ensures the implementation of the action plan. Step 5 (monitor, evaluate, and course correct) aims to ensure that the lead ministry monitors the action plan for each community setting, evaluates the impact of the plan, and coordinates cyclical review and analysis (29). Step 6 (cross-cutting themes) refers to topics such as equity, gender, inclusion, and non-discrimination, that should be integrated throughout the five previous steps, rather than addressed as stand-alone topics. The specific parameters for each step are defined in the Supplementary Information (Table S5).

**Figure 1.**
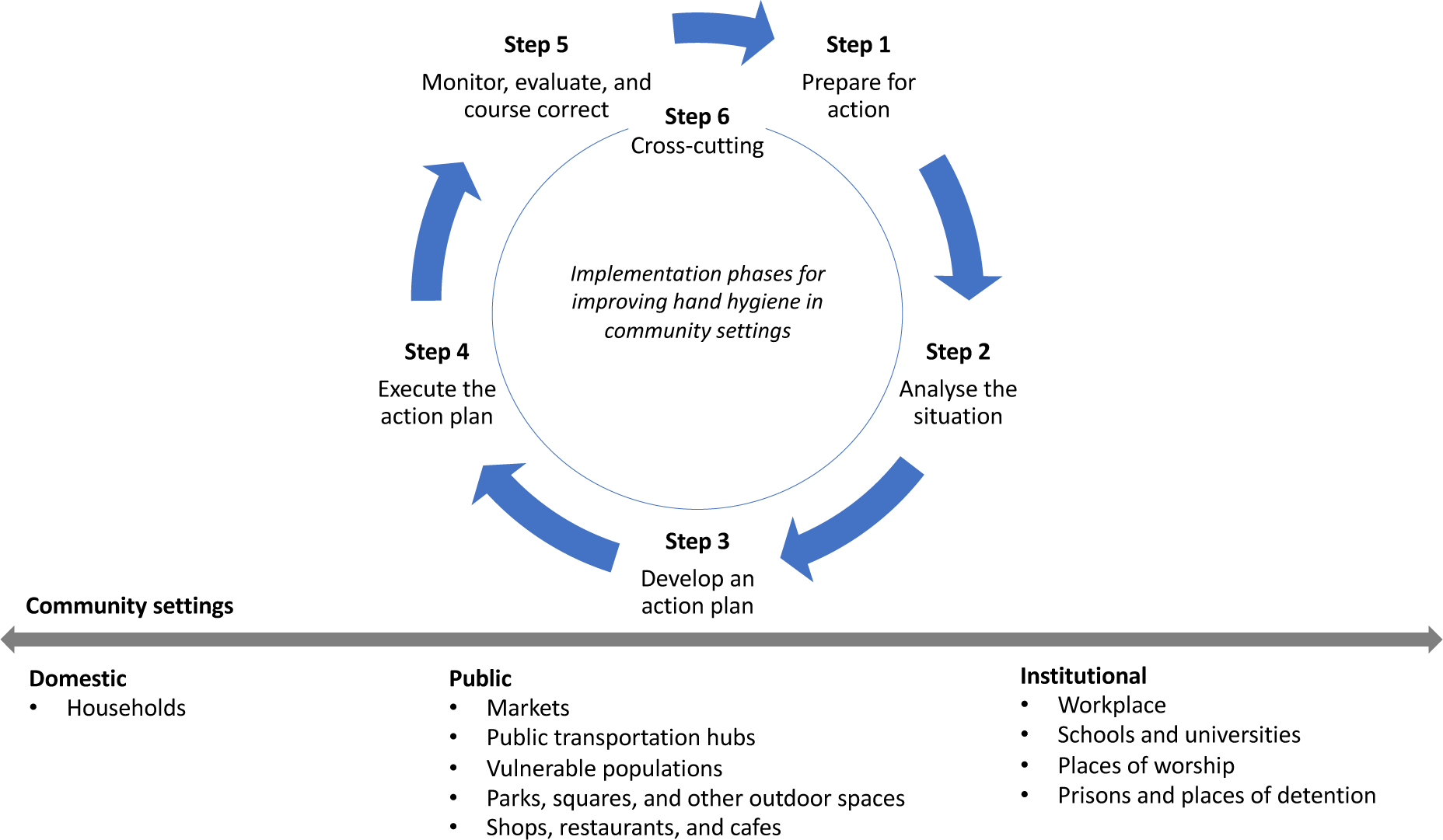
Conceptual framework developed for this scoping review that includes a six-step approach to support the implementation of recommendations for hand hygiene in community settings. The framework was adapted from the 5-step approach from Burke et al. (2012) (26) and WHO (2009) (27) to include a cross-cutting sixth step.

### Identifying relevant studies (stage 2)

The search strategy queried (1) Google search engine, (2) websites of international organisations known to work on hand hygiene (Table S2), and 3) (Healthcare Management Information Consortium) (HMIC), a grey literature database. The electronic database search was conducted using keyword searches and MeSH terms (Table S3). Each search term, with synonyms, variations, and subject headings, was combined and truncated to capture all possible variations of relevant terms. The search in Google was carried out using the anonymous function in the web browser (Chrome) to reduce the influence of the reviewer’s (CM) individual search history. Search strings were constructed by using multiple combinations of search terms from Supplementary Information Table S3. For each combination of search terms, the first 10 pages of Google were reviewed by one reviewer (CM) (30). The reference lists of included documents were also hand-searched for any additional relevant documents. The search was limited to English and publication date was restricted to 1 January 1990 onwards to capture current tools and implementation guidance (31).

### Study selection (stage 3)

Documents were included if they met all of the following criteria: (1) international tool or guidance document, (2) is relevant to one of the six implementation steps in the conceptual framework, (3) targets at least one community setting, as defined in the conceptual framework, (4) published by an international non-governmental organisation (NGO), multilateral agency or public health agency, (5) published in English, and (6) published between 1 January 1990 and 1 August 2023.

Training handbooks and training modules related to hand hygiene were excluded from this review as they are intended for teaching and learning step-by-step procedures. Tools published as an application software were also excluded. In addition, we excluded tools for humanitarian settings as internationally agreed guidance on hand hygiene in humanitarian settings or complex emergencies is available through the Sphere standards for water, sanitation, and hygiene (WASH) promotion (32). Implementation guidance documents and tools that have a more recent edition available and country- or region-specific implementation tools were excluded as they do not support the global implementation of recommendations for hand hygiene.

All documents retrieved from electronic searches were transferred to EndNote. Screening was completed in two stages: (1) title and document objective were screened for eligibility by one reviewer (CM); and (2) full texts of all potentially eligible documents were retrieved and independently assessed for inclusion by two reviewers (CM and LB). Disagreement between reviewers on inclusion was resolved through arbitration by a third reviewer (OC).

### Charting the data (stage 4)

Tool characteristics and implementation recommendations from included documents were independently double extracted by two reviewers (CM and LB) using a standardised data extraction template in MS Excel (Table S4) and then cross-checked for accuracy. As with inclusion, a third reviewer (OC) provided arbitration if agreement on extraction could not be reached. The data extraction form included information on document characteristics, such as author, year of publication, target setting, as well as on specific parameters related to the six implementation steps for hand hygiene in community settings described in the conceptual framework (figure 1). Definitions of the parameters are included in the Supplementary Information (Table S5). Implementation guidance for each parameter was extracted from included documents where possible.

### Collating, summarising, and reporting the results (stage 5)

To synthesise evidence, implementation guidance and tools were first mapped against the six implementation steps, then summarised for each implementation step. We also summarised the number of documents that provided guidance or tools across all six steps. Finally, we identified steps with little or no implementation guidance or tools.

## Results

### Search results

Electronic searches were conducted in August 2023, identifying 2,990 records (2,000 from Google, 957 from organisation websites, and 33 from reference screening). No documents were retrieved from the grey literature database. 100 documents were sought for retrieval for full-text screening. Finally, 35 documents with implementation guidance and tools are included in the review (Figure 2). The 65 documents excluded during full-text review are listed in the Supplementary Information (Table S6) with reasons for exclusion.

**Figure 2.**
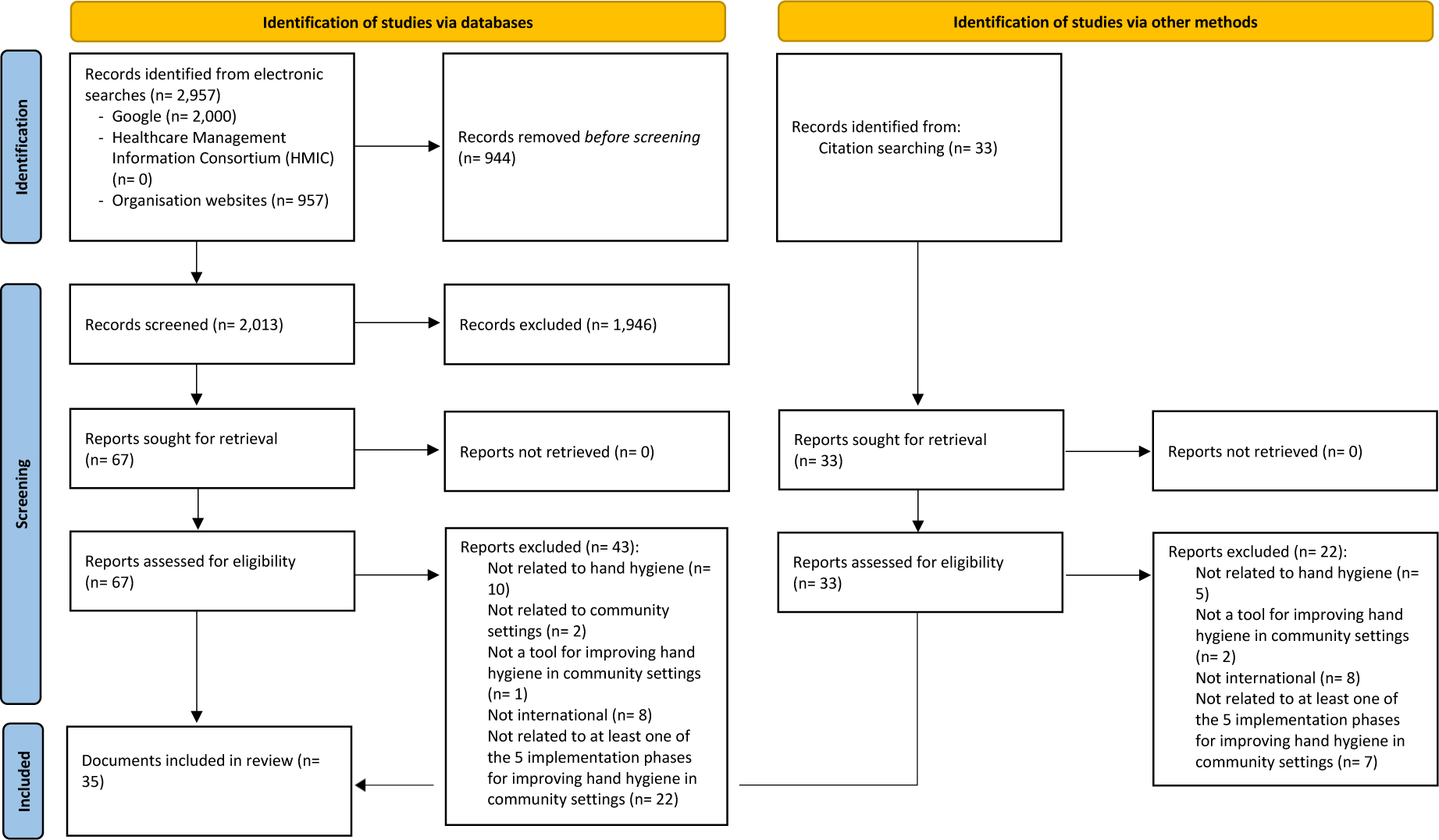
PRISMA flow diagram.

### Description of included studies

We included 35 documents in the review, consisting of 30 implementation guidance documents and 5 stand-alone tools. Among the 30 implementation guidance documents, 22 provide implementation guidance only and 8 include implementation guidance and at least one tool (a total of 16 tools are extracted from the 8 documents with tools) (Figure 3). Among these implementation guidance documents, we extracted 207 implementation recommendations (Figure 3). 162 implementation recommendations were extracted from the 22 implementation guidance documents and 45 recommendations were extracted from the 8 documents with implementation guidance and tools. We also included a total of 21 tools in the review, including 5 stand-alone tools and 16 tools extracted from 8 documents with implementation guidance and tools.

**Figure 3.**
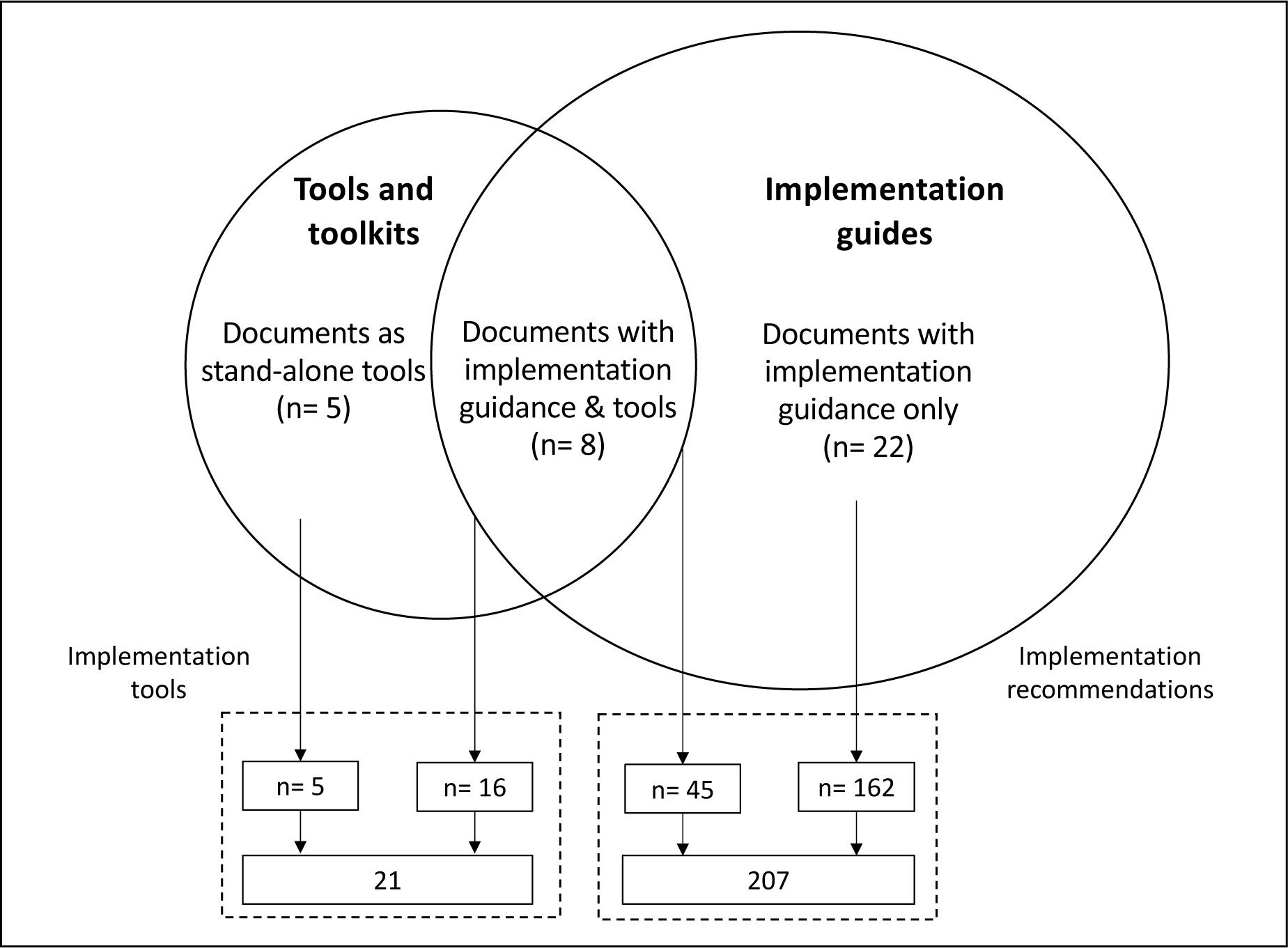
Diagram of included documents that are categorised as tools (n= 5) or implementation guidance documents (n= 30). Of the 30 implementation guidance documents, 22 include implementation guidance only and 8 include implementation guidance and at least one tool (a total of 16 tools are extracted from the 8 documents with tools). We identified 5 documents as stand-alone tools.

48% percent of included documents are published by multilateral agencies (WHO, UNICEF), 34% by international NGOs, 9% by global partnerships (e.g., Global Handwashing Partnership), 6% by academic institutions, and 3% by development agencies. Most included documents are WASH-related (77%, n= 27), while 23% (n= 8) are hand hygiene specific. There are no documents with a broader aim that provide implementation guidance or tools on hand hygiene (e.g., cholera guidance that include hand hygiene tools). Most documents are for the programme level (66%, n= 23), while 34% (n= 12) are for national-level planning and implementation. Documents for the programme level are typically widely applicable to any programme related to hand hygiene, though the applicability may vary by community setting. Similarly, national-level documents are applicable widely across countries. Almost half of included documents target the institutional setting (40%, n= 14), one targets the domestic setting (3%), and none target the public setting. The remaining documents do not specify the targeted community setting (57%, n= 20). Among documents for the institutional setting, 12 concern schools and 2 concern the workplace. The included documents are for high-, medium-, and low-income settings and applicable across these settings. All documents were published between 2005 and 2023. Full details of included documents can be found in the Supplementary Information (Table S7).

### Implementation guidance and tools for hand hygiene in community settings

There is guidance for the implementation of hand hygiene recommendations in community settings, but there is a limited number of available tools (Figure 4). The 207 implementation recommendations and 21 tools were mapped against the six implementation steps for hand hygiene in community settings. Most guidance and tools are for the middle and end of the implementation cycle. Specifically, these are for step 5 (monitoring, evaluation, and course correction) (27%, 62/228), followed by step 2 (analyse the situation) (23%, 52/228), step 3 (develop an action plan) (22%, 50/228), and step 1 (prepare for action) (16%, 37/228) (Figure 4). There is little guidance and tools for step 4 (execute the action plan) (7%, 16/228) and step 6 (cross-cutting) (5%, 11/228). Only five documents provided guidance or a tool across the first five implementation steps. Of these documents, only two also provided guidance or a tool for the sixth implementation (cross-cutting) (Table S8).

**Figure 4.**
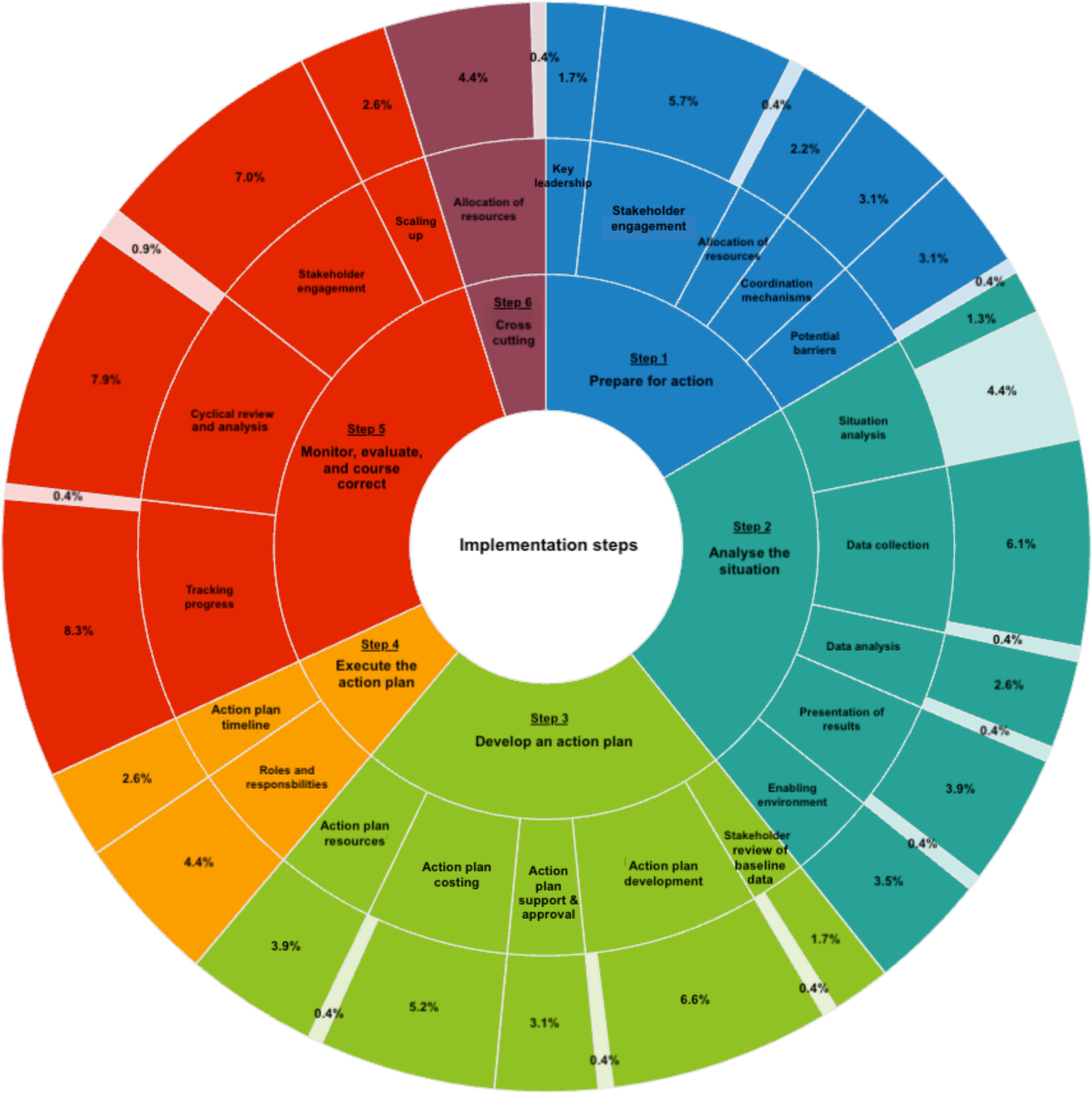
Sunburst diagram of implementation guidance documents and toolkits for the six implementation steps, in addition to cross-cutting issues. For each key parameter, light shading indicates tools and dark shading indicates implementation guidance.

### Implementation guidance for hand hygiene in community settings

The 207 implementation recommendations were mapped across key parameters for the six implementation steps (Table 1). Overall, most implementation guidance was for ‘tracking progress’ and ‘cyclical review and analysis’ (n= 19 and 18, respectively), both part of step 5. The pieces of guidance ranged from 3 to 16 for all other steps. The least guidance was provided for ‘identifying situation analysis tools’ (step 2, n= 3), ‘key leadership’ (step 1, n= 4) and ‘stakeholder review of baseline data’ (step 3, n= 4).

**Table 1.** Implementation guidance for hand hygiene in community settings.

| Implementation phase | Key parameter | Implementation guidance (% , n) | References |
| --- | --- | --- | --- |
| **Step 1:** prepare for action | Key leadership | 2% (4) | IRC, 2007 (33); UNICEF, 2016 (34); UNICEF, 2019 (35); UNICEF, 2022 (36) |
|  | Stakeholder engagement | 6% (12) | World Bank, 2005 (37); World Bank & UNICEF, 2005 (38); IRC, 2007 (33); Live & Learn, 2011 (39); UNICEF, 2012 (40); UNICEF, 2016 (34); Rotary International, 2017a (41); Rotary International 2017c (42); WaterAid, 2017 (43); GHP, 2018 (44); UNICEF, 2019 (35); UNICEF, 2022 (36) |
|  | Allocation of resources for planning | 2% (5) | World Bank, 2005 (37); IRC, 2007 (33); UNICEF, 2012 (40); UNICEF, 2019 (35); UNICEF, 2022 (36) |
|  | Coordination mechanisms | 3% (7) | World Bank, 2005 (37); IRC, 2007 (33); UNICEF, 2012 (40); Rotary International, 2017a (41); Rotary International 2017c (42); UNICEF, 2019 (35); UNICEF, 2022 (36) |
|  | Potential barriers and challenges | 3% (7) | UNICEF, 2012 (40); UNICEF, 2016 (34); Rotary International, 2017a (41); UNICEF, 2017 (43); WaterAid, 2017 (43); UNICEF, 2021 (45); World Vision, 2021 (46) |
| **Step 2:** analyse the situation | Identification of tools for situation analysis | 1% (3) | Rotary International, 2017a (41); Concern Worldwide, 2021 (47); World Vision, 2021 (46) |
|  | Data collection | 7% (14) | World Bank, 2005 (37); IRC, 2007 (33); WBCSD, 2014 (48); PPPHW, 2015 (49); EAWAG, 2016 (50); UNICEF, 2016 (34); LSHTM, 2017 (51); Rotary International, 2017b (52); WaterAid, 2017 (43); UNICEF, 2019 (35); Concern Worldwide, 2021 (47); UNICEF, 2021 (45); World Vision, 2021 (46); World Vision, 2022 (53) |
|  | Data analysis | 3% (6) | PPPHW, 2015 (49); UNICEF & WHO, 2016 (54); Rotary International, 2017a (41); UNICEF, 2021 (45); WaterAid, 2017 (43); UNICEF, 2022 (36) |
|  | Presentation of results | 4% (9) | WBCSD, 2014 (48); PPPHW, 2015 (49); UNICEF & WHO, 2016 (54); LSHTM, 2017 (51); Rotary International, 2017a (41); WaterAid, 2017 (43); UNICEF, 2019 (35); UNICEF, 2021 (45); UNICEF, 2022 (36) |
|  | Enabling environment | 4% (8) | IRC, 2007 (33); UNICEF, 2016 (55); UNICEF, 2016b (55); LSHTM, 2017 (51); |
|  |  |  | Rotary International, 2017a (41); Rotary International, 2017c (42); SWA, 2020 (56); World Vision, 2022 (53) |
| **Step 3: develop an action plan** | Stakeholder review of baseline data | 2% (4) | World Bank, 2005 (37); IRC, 2007 (33); UNICEF, 2012 (40); UNICEF, 2022 (36) |
|  | Action plan development | 7% (15) | World Bank, 2005 (37); World Bank & UNICEF, 2005 (38); IRC, 2007 (33); Live & Learn, 2011 (39); GIZ, 2013 (57); WBCSD, 2014 (48); UNICEF, 2016 (34); LSHTM, 2017 (51); Rotary International, 2017a (41); Rotary International, 2017c (42); GHP, 2018 (44); UNICEF, 2019 (35); Concern Worldwide, 2021 (47); UNICEF, 2022 (36); World Vision, 2021 (46) |
|  | Action plan support and approval | 3% (7) | World Bank, 2005 (37); IRC, 2007 (33); UNICEF, 2012 (40); Rotary International, 2017a (41); UNICEF, 2019 (35); UNICEF, 2022 (36); World Vision, 2022 (53) |
|  | Action plan costing | 6% (12) | World Bank, 2005 (37); World Bank & UNICEF, 2005 (38); IRC, 2007 (33); UNICEF, 2012 (40); GIZ, 2013 (57); UNICEF, 2016 (34); Rotary International, 2017a (41); Rotary International, 2017c (42); UNICEF, 2019 (35); Concern Worldwide, 2021 (47); UNICEF, 2022 (36); World Vision, 2022 (53) |
|  | Action plan resources | 4% (9) | World Bank & UNICEF, 2005 (38); IRC, 2007 (33); GIZ, 2013 (57); LSHTM, 2017 (51); Rotary International, 2017a (41); Rotary International, 2017c (42); SWA, 2020 (56); UNICEF, 2022 (36); World Vision, 2022 (53) |
| **Step 4: execute plans** | Roles and responsibilities | 5% (10) | World Bank, 2005 (37); World Bank & UNICEF, 2005 (38); IRC, 2007 (33); UNICEF, 2012 (40); GIZ, 2013 (57); UNICEF, 2016 (34); LSHTM, 2017 (51); Rotary International, 2017c (42); UNICEF, 2022 (36); World Vision, 2022 (53) |
|  | Action plan completion timeline | 3% (6) | World Bank, 2005 (37); IRC, 2007 (33); UNICEF, 2016 (34); Rotary International, 2017c (42); UNICEF, 2019 (35); UNICEF, 2022 (36) |
| **Step 5: monitor, evaluate, and course correct** | Tracking progress | 9% (19) | World Bank, 2005 (37); IRC, 2007 (33); World Bank, 2010 (58); Live & Learn, 2011 (39); UNICEF, 2011 (59); UNICEF, 2012 (40); UNICEF, 2013 (60); WBCSD, 2014 (48); PPPHW, 2015 (49); UNICEF, 2016 (34); LSHTM, 2017 (51); Rotary International, 2017a (41); UN-Water, 2017 (61); GHP, 2018 (44); UNICEF, 2019b (62); Concern Worldwide, 2021 |
|  |  |  | (47); UNICEF, 2021 (45); World Vision, 2021 (46); UNICEF, 2022 (36) |
|  | Cyclical review and analysis | 9% (18) | World Bank, 2005 (37); IRC, 2007 (33); Live & Learn, 2011 (39); UNICEF, 2011 (59); UNICEF, 2012 (40); UNICEF, 2013 (60); UNICEF, 2016a (34); UNICEF, 2016b (55); UNICEF & WHO, 2016 (54); LSHTM, 2017 (51); Rotary International, 2017a (41); WaterAid, 2017 (43); UNICEF, 2019 (35); UN-Water, 2017 (61); Concern Worldwide, 2021 (47); UNICEF, 2021 (45); World Vision, 2021 (46); UNICEF, 2022 (36) |
|  | Stakeholder engagement | 8% (16) | World Bank & UNICEF, 2005 (38); IRC, 2007 (33); Live & Learn, 2011 (39); UNICEF, 2011 (59); UNICEF, 2012 (40); UNICEF, 2013 (60); WBCSD, 2014 (48); UNICEF, 2016 (34); LSHTM, 2017 (51); Rotary International, 2017a (41); WaterAid, 2017 (43); UNICEF, 2019 (35); Concern Worldwide, 2021 (47); UNICEF, 2021 (45); World Vision, 2021 (46); World Vision, 2022 (53) |
|  | Scaling up | 3% (6) | IRC, 2007 (33); Live & Learn, 2011 (39); UNICEF, 2012 (40); UNICEF, 2016 (55); World Vision, 2021 (46); UNICEF, 2022 (36) |
| **Step 6: Cross-cutting** | Equity, gender, inclusion, and non-discrimination | 5% (10) | World Bank & UNICEF, 2005 (38); IRC, 2007 (33); UNICEF, 2012 (40); PPPHW, 2015 (49); Rotary International, 2017a (41); WaterAid, 2017 (43); UNICEF, 2019 (35); Concern Worldwide, 2021 (47); UNICEF, 2021 (45); World Vision, 2022 (53) |
| **Total** |  | **100% (207)** | **30 documents** |

### Tools for hand hygiene in community settings

In total, we included 21 individual tools (5 stand-alone and 16 tools extracted from 8 implementation guidance documents). Most tools are for step 2 (analysing the situation) (57%, 12/21). Of these, most are for undertaking a situation analysis (75%, 9/12) (Table 2). Other tools were commonly mapped to step 3 (develop an action plan) (14%, 3/21) and step 5 (monitoring, evaluating, and course correcting) (14%, 3/21). There is only one tool for step 1 (prepare for action) and for step 6 (cross-cutting issues). There are no tools available for step 4 (executing the action plan). The tools ranged from checklists to surveys, self-assessment tools, planning and costing tools, and a list of monitoring questions (Table 3). Of the 5 stand-alone tools, only one is specific to hand hygiene only, which is to cost interventions for improving hand hygiene in the domestic setting (63). The other 4 stand-alone tools are part a wider tool relevant to WASH (64–67).

**Table 2.** Tools to support the implementation of hand hygiene recommendations in community settings (one document can contain multiple tools)

| Implementation phase | Key actions | # of tools | Relevant tool(s) | Objective | References |
| --- | --- | --- | --- | --- | --- |
| **Step 1:** prepare for action | Stakeholder engagement | 1 | Tool to advocate for hygiene | To make the case for hand hygiene | (PPPHW, 2015) (49) |
|  | Potential barriers and challenges | 3 | SWOT analysis | To identify the strengths, weaknesses, opportunities, and threats (SWOT) for a handwashing programme | (World Bank, 2005) (37) |
|  |  |  | SWOT external analysis | To identify external factors that may influence the handwashing programme | (World Bank, 2005) (37) |
|  |  |  | WASH Bottleneck Analysis Tool (BAT) | To help formulate costed and prioritized Action Plans to remove the bottlenecks that constrain the WASH sector and hinder the delivery of sustainable WASH services. | (UNICEF, 2011) (67) |
| **Step 2:** analyse the situation | Identification of tools for situation analysis | 16 | SWOT external analysis | To identify external factors that may influence the handwashing programme | (World Bank, 2005) (37) |
|  |  |  | School WASH survey (page 24) | To conduct a thorough needs assessment of a school for improving school WASH conditions | (Peace Corps, 2017) (66) |
|  |  |  | Barrier analysis tool | To identify the barriers that prevent people experiencing marginalisation from accessing WASH on an equal basis with others | (WaterAid, 2017) (43) |
|  |  |  | Gender analysis tool | To understand how a particular situation affects women and men differently | (WaterAid, 2017) (43) |
|  |  |  | Stakeholder analysis tool | To map current stakeholders and assess the role they play, particularly in helping you to achieve inclusive WASH | (WaterAid, 2017) (43) |
|  |  |  | Power analysis tool | To identify who has formal power over an issue, who has informal power, and who can influence those with power | (WaterAid, 2017) (43) |
|  |  |  | Disability self-assessment tool | To identify specific actions that can be taken to increase the inclusion of disabled people in programmes | (WaterAid, 2017) (43) |
|  |  |  | Political economy analysis tool | To provide a structured approach for analysing how change happens, from the national to the local level | (WaterAid, 2017) (43) |
|  |  |  | WASH assessment tool: self-assessment & employee survey | To measure the current WASH status of an organisation or business | (WaterAid, 2021) (65) |
|  |  |  | Tool for identifying persons with disabilities | To identify persons with disabilities and disaggregate WASH data by disability | (UNICEF, 2021) (45) |
|  |  |  | Self-assessment tool to understand current practices | To understand the current level of WASH provisions business-wide | (WBCSD, 2014) (48) |
|  |  |  | Core hygiene questions | To support increased monitoring of hand hygiene in schools | (UNICEF & WHO, 2016) (54) |
|  |  |  | Observation checklist for WASH in schools | To understand observed existing WASH infrastructure at the school | (Rotary International, 2017a) (52) |
|  |  |  | School WASH survey | To understand WASH practices and education at the school | (Rotary International, 2017a) (52) |
|  |  |  | Focus group discussion questions | To understand the challenges regarding WASH at the school | (Rotary International, 2017a) (52) |
|  |  |  | WASH in schools target challenge measurements | To gather baseline measurements on WASH in schools targets | (Rotary International, 2017a) (52) |
|  | Data collection | 1 | WASH assessment tool (tab 3 and 5b) | To measure the current WASH status of an organisation or business | (WaterAid, 2021) (65) |
|  | Data analysis | 1 | WASH assessment tool (tabs 4 and 5d) | To measure the current WASH status of an organisation or business | (WaterAid, 2021) (65) |
|  | Presentation of results | 1 | WASH assessment tool (tabs 4 and 5d) | To measure the current WASH status of an organisation or business | (WaterAid, 2021) (65) |
| **Step 3:** develop an action plan | Stakeholder review of baseline data | 1 | WASH assessment tool (tab 5a) | To measure the current WASH status of an organisation or business | (WaterAid, 2021) (65) |
|  | Action plan development | 1 | WASH assessment tool (tab 6) | To measure the current WASH status of an organisation or business | (WaterAid, 2021) (65) |
|  | Action plan costing | 1 | Costing tool for estimating the cost of interventions to improve hand hygiene in the domestic settings | This tool aims to provide country-specific cost estimates for achieving universal hand hygiene in households by 2030 | (WHO and UNICEF, 2021) (68) |
| **Step 5:** monitor, evaluate, and course correct | Tracking progress | 2 | Participation ladder | To monitor participation in a programme over time | (WaterAid, 2017) (43) |
|  |  |  | Washington Group questions for collecting data on disability | A set of questions designed to identify people with a disability, such as difficulty performing basic universal activities such as walking, seeing, hearing, cognition, self-care, and communication | (WaterAid, 2017) (43) |
|  | Cyclical review and analysis | 2 | Hand hygiene acceleration framework tool | To track the process that a government has undergone to develop and implement a plan of action for hand hygiene improvement and assesses the quality of that plan | (WHO, UNICEF, and WaterAid, 2023) (64) |
|  |  |  | WASH assessment tool | To measure the current WASH status of an organisation or business | (WaterAid, 2017) (43) |
| **Step 6:** cross-cutting | Equity, gender, inclusion, and non-discrimination | 1 | Questions on accessibility for children with physical disabilities | To assess the accessibility of facilities and hygiene education programmes for children with physical disabilities | (UNICEF, 2011) (59) |

## Discussion

This scoping review includes 35 documents, comprising 30 implementation guidance documents and 5 stand-alone tools published between 2005 and 2023. Most documents are for all community settings, suggesting these are intended to be generalisable across the different community settings. The only exception is that 12 documents focused exclusively on school settings. In addition, the included documents are for high-, medium-, and low-income settings and applicable across country settings. While the included documents have a global focus, most implementation guidance and tools will require some form of adaptation to specific country contexts where implementation modalities and options can vary significantly. Most documents were published by multilateral agencies, such as UNICEF and WHO, and NGOs, such as WaterAid and World Vision, that have experience designing and implementing WASH or hand hygiene programmes.

Across the 35 included documents, we identified a total of 21 tools and 207 implementation recommendations, highlighting a discrepancy between available implementation guidance and tools to accompany the guidance. The 21 tools include 5 stand-alone tools and 16 tools embedded within broader WASH or hand hygiene guidance documents. The implementation guidance and tools were mapped to a six-step implementation conceptual framework for hand hygiene in community settings, which was adapted from an existing one that was also used by WHO for the hand hygiene in healthcare settings guidelines. WHO also used a similar six-step approach for the sustainable implementation of national action plans on antimicrobial resistance (69). However, no other review has mapped implementation guidance or tools to the 5-step approach adapted for this review.

The amount of available implementation guidance varied across the implementation steps, as well as amongst parameters within each step. Most implementation guidance was mapped to steps 1 (prepare for action), 2 (analyse the situation), 3 (develop an action plan), and 5 (monitor, evaluate, and course correct) of the conceptual framework, with limited guidance for step 4 (executing the action plan) and step 6 (cross-cutting issues). This might be expected, as each implementation step may not require the same amount of guidance. For example, more guidance for steps 1-3 and 5, compared to step 4, is expected as it relates to supporting end-users to prepare an action plan and monitor its implementation. However, for steps 1-3 and 5, where guidance is available, more guidance may also be required for specific parameters, such as ‘identifying key leadership’ and ‘allocation of resources for planning’ under step 1, ‘data analysis’ and ‘enabling environment’ under step 2, ‘stakeholder review of baseline data’ under step 3, and ‘scaling up’ under step 5. Further implementation guidance is needed for step 6 (cross-cutting themes). While cross-cutting themes were classified under step 6 of the implementation framework, in practice they should be integrated across the previous five steps to enhance equitable guideline uptake. In addition, this scoping review highlights the fragmentation of guidance and tools across the implementation cycle. Only two included documents provided guidance across the six steps, suggesting that guidance among each included document is rarely comprehensive.

The limited number of relevant tools suggests that more comprehensive or implementation step-specific tools may be needed to support the implementation of hand hygiene recommendations. Only 1 tool of the 5 stand-alone tools was developed for hand hygiene specifically, further suggesting the limited number of tools tailored for hand hygiene in community settings. Like implementation guidance, the tools were inconsistently available across the implementation steps. Over half of identified tools are for step 2 (analyse the situation) and primarily for one sub-step (undertaking a situation analysis). The identified tools varied in terms of content and purpose and can support various data collection and decision-making efforts for a more systematic and tailored adoption of guideline recommendations. However, more tools are needed to effectively be used as a guideline knowledge translation strategy to facilitate the implementation of recommendations into action, such as tools to support the costing of national action plans. In addition, further research can assess whether and how the tools translate into the implementation of recommendations. Tools can act as an important link between guideline recommendations and the development of national policies and programmes for hand hygiene in community settings. This is especially crucial for moving from emergency response, such as during the COVID-19 pandemic, to building sustainable and equitable services (71).

Despite gaps, the identified tools and implementation guidance can be used for the uptake of guidelines for hand hygiene in community settings to further contextualise and adapt recommendations. Resources have been published in other public health areas for the implementation of guideline recommendations. For example, WHO published a set of documents to support the implementation of recommendations for the hand hygiene in health care guidelines (72). These resources are intended to guide healthcare facilities to develop action plans for hand hygiene. Similarly, WHO published implementation guidance to advise countries on how to develop and implement national malaria strategic plans (69), recommended as part of the guidelines for malaria (73). While gaps remain, the implementation guidance and tools identified in this review can be leveraged to support the upcoming WHO guidelines on hand hygiene in community settings.

## Limitations

This scoping review has several limitations. First, most documents were retrieved through the Google search engine and the websites of organisations known to work on hand hygiene. Implementation guidance documents and tools may have been missed if the publishing organisation’s website was not searched or the content was not indexed by Google. In addition, the one database that was searched for grey literature did not provide any relevant results. Second, despite a comprehensive search strategy, relevant documents may not have appeared in the search results due to the lack of consistency in terminology for tools and implementation guidance amongst organisations. Third, the search was restricted to English and may have missed relevant documents published in other languages. Fourth, we excluded tools and guidance documents published by governments, which can also be used for implementing hand hygiene in specific regional or national contexts. Fourth, the analysis did not seek to comparatively and qualitatively assess the included tools and implementation guidance. Instead, specific guidance was mapped to the implementation steps to understand where guidance is available and where there are gaps. Fourth, implementation guidance and tools were not disaggregated by community settings as most guidance and tools were for all community settings, thereby limiting the use of guidance and tools for community-specific settings. Finally, we did not undertake a quality assessment of the tools nor assess whether implementation guidance was cited by evidence.

## Conclusion

Overall, this scoping review identified 35 documents with both tools and implementation guidance for hand hygiene in community settings. The mapping exercise suggests that implementation guidance is available, yet inconsistently available for the different implementation steps. In addition, few implementation tools exist to support the more general guidance. Future work should focus on developing comprehensive practical tools for the implementation of hand hygiene recommendations in community settings, in line with international guidelines.

## Data Availability

All data produced in the present study are available upon reasonable request to the authors

## Author contributions

CM, LB, BAC, CC, KC, JC, RD, REN, AT, BG, JEM and OC informed the study protocol. CM carried out the database and grey literature search with input from LB, OC, and JEM. CM and LB screened the retrieved articles for inclusion and extracted the data with input from OC and JEM. CM led the data synthesis, while CM, LB, OC and JEM led the presentation of results with inputs from co-authors. CM wrote the first draft of the manuscript and all co-authors provided inputs on subsequent drafts. LB, OC, and JEM provided overall supervision, leadership and advice. CM is the guarantor.

## Author approval

All authors have seen and approved the manuscript for publication.

## Data availability

Data are available upon reasonable request.

