## Supporting Information for "Current international tools and guidance for the implementation of hand hygiene recommendations in community settings: a scoping review"

**Table S1**. The completed Preferred Reporting Items for Systematic reviews and Meta-Analyses extension for Scoping Reviews (PRISMA-ScR) checklist (74).

| SECTION | ITEM | PRISMA-ScR CHECKLIST ITEM | REPORTED ON PAGE # |
|---|---|---|---|
| **TITLE** | | | |
| Title | 1 | Identify the report as a scoping review. | 1 |
| **ABSTRACT** | | | |
| Structured summary | 2 | Provide a structured summary that includes (as applicable): background, objectives, eligibility criteria, sources of evidence, charting methods, results, and conclusions that relate to the review questions and objectives. | 2 |
| **INTRODUCTION** | | | |
| Rationale | 3 | Describe the rationale for the review in the context of what is already known. Explain why the review questions/objectives lend themselves to a scoping review approach. | 3 – 4 |
| Objectives | 4 | Provide an explicit statement of the questions and objectives being addressed with reference to their key elements (e.g., population or participants, concepts, and context) or other relevant key elements used to conceptualize the review questions and/or objectives. | 4 |
| **METHODS** | | | |
| Protocol and registration | 5 | Indicate whether a review protocol exists; state if and where it can be accessed (e.g., a Web address); and if available, provide registration information, including the registration number. | 4 |
| Eligibility criteria | 6 | Specify characteristics of the sources of evidence used as eligibility criteria (e.g., years considered, language, and publication status), and provide a rationale. | 8 |
| Information sources | 7 | Describe all information sources in the search (e.g., databases with dates of coverage and contact with authors to identify additional sources), as well as the date the most recent search was executed. | 7 |
| Search | 8 | Present the full electronic search strategy for at least 1 database, including any limits used, such that it could be repeated. | Table S3 |
| Selection of sources of evidence | 9 | State the process for selecting sources of evidence (i.e., screening and eligibility) included in the scoping review. | 5 |
| Data charting process | 10 | Describe the methods of charting data from the included sources of evidence (e.g., calibrated forms or forms that have been tested by the team before their use, and whether data charting was done independently or in duplicate) and any processes for obtaining and confirming data from investigators. | 8 – 9 |
| Data items | 11 | List and define all variables for which data were sought and any assumptions and simplifications made. | Table S4 |
| Critical appraisal of individual sources of evidence | 12 | If done, provide a rationale for conducting a critical appraisal of included sources of evidence; describe the methods used and how this information was used in any data synthesis (if appropriate). | n/a |
| Synthesis of results | 13 | Describe the methods of handling and summarizing the data that were charted. | 9 |

| SECTION | ITEM | PRISMA-ScR CHECKLIST ITEM | REPORTED ON PAGE # |
|---|---|---|---|
| **RESULTS** | | | |
| Selection of sources of evidence | 14 | Give numbers of sources of evidence screened, assessed for eligibility, and included in the review, with reasons for exclusions at each stage, ideally using a flow diagram. | 9 |
| Characteristics of sources of evidence | 15 | For each source of evidence, present characteristics for which data were charted and provide the citations. | Table S7 |
| Critical appraisal within sources of evidence | 16 | If done, present data on critical appraisal of included sources of evidence (see item 12). | n/a |
| Results of individual sources of evidence | 17 | For each included source of evidence, present the relevant data that were charted that relate to the review questions and objectives. | 12 – 20 |
| Synthesis of results | 18 | Summarize and/or present the charting results as they relate to the review questions and objectives. | 12 – 20 |
| **DISCUSSION** | | | |
| Summary of evidence | 19 | Summarize the main results (including an overview of concepts, themes, and types of evidence available), link to the review questions and objectives, and consider the relevance to key groups. | 21 – 22 |
| Limitations | 20 | Discuss the limitations of the scoping review process. | 23 |
| Conclusions | 21 | Provide a general interpretation of the results with respect to the review questions and objectives, as well as potential implications and/or next steps. | 23 |
| **FUNDING** | | | |
| Funding | 22 | Describe sources of funding for the included sources of evidence, as well as sources of funding for the scoping review. Describe the role of the funders of the scoping review. | 24 |

**Table S2**. List of websites of international organisations known to work on hand hygiene searched.

| Agency or resource hub | Website |
|---|---|
| Global Handwashing Partnership | https://globalhandwashing.org/ |
| Global WASH Cluster | www.washcluster.org |
| International Red Cross and Red Crescent (ICRC) | www.icrc.org |
| International Federation of the Red Cross (IFRC) | www.ifrc.org |
| International Labour Organization (ILO) | https://www.ilo.org |
| IRC WASH | https://www.ircwash.org/ |
| Oxfam | www.oxfam.org.uk |
| Sanitation Learning Hub | https://sanitationlearninghub.org/ |
| Save the Children | www.nrc.no |
| SNV Netherlands Development Organisation | https://snv.org/ |
| Sustainable Sanitation Alliance | https://www.susana.org/en/ |
| United Nations Human Settlement Programme (UN Habitat) | https://unhabitat.org/ |
| United Nations High Commissioner for Refugees (UNHCR) | www.unhcr.org |
| United Nations Children's Fund (UNICEF) | www.unicef.org |
| US Centers for Disease Control and Prevention (CDC) | www.cdc.gov |
| WASH'Em | https://www.washem.info/ |
| WaterAid | https://www.wateraid.org/us/ |
| World Bank | https://www.worldbank.org/en/home |
| Water, Education and Development Centre (WEDC) | www.wedc.lboro.ac.uk |
| Water and Sanitation for the Urban Poor (WSUP) | https://www.wsup.com/ |
| Deutsche Gesellschaft für Internationale Zusammenarbeit (GIZ) | https://www.giz.de |
| United States Agency for International Development (USAID) | https://www.usaid.gov/ |

**Table S3**. Search strategy.

| Concept | Number | Search terms |
|---|---|---|
| Hand hygiene | 1 | handwash* OR hand wash* OR hygiene* OR hand clean* OR soap* OR clean hand* OR HWWS OR ABHR OR hand sanit* OR hand rub OR hand disinfect* |
| Community settings | 2 | communit* OR household* OR domestic OR public space* OR public setting* OR institution* OR work* OR occupation* OR school* OR prison* OR market* OR transport* OR relig* |
| Implementation guidance and tools | 3 | implementation guide OR implementation guides OR practical OR guide OR practical guides OR operational guide OR operational guides OR handbook* OR manual* OR tool* OR tool kit |
| | 4 | 1 AND 2 AND 3 |

**Table S4**. Data extraction template.

| **Document** | | | **General** | | | | | **Step 1: prepare for action** | | | | | **Step 2: analyse the situation** | | |
|---|---|---|---|---|---|---|---|---|---|---|---|---|---|---|---|
| Author | Title | Year | Community setting | Target user | Target level | WASH vs HH | Objective | Key leadership | Stakeholder engagement | Allocation of resources for planning | Coordination mechanisms | Potential barriers and challenges | Identification of tools for situation analysis | Undertake the situation analysis | Data collection |

(data extraction template continued)

| **Step 2 (continued)** | | | **Step 3: develop an action plan** | | | | **Step 4: execute plans** | | | | **Step 5: monitor, evaluate, and course correct** | | **Step 6: cross-cutting** |
|---|---|---|---|---|---|---|---|---|---|---|---|---|---|
| Data analysis | Presentation of results | Enabling environment | Stakeholder review of baseline data | Action plan development | Action plan support and approval | Action plan costing | Roles and responsibilities | Action plan completion timeline | Tracking progress | Cyclical review and analysis | Stakeholder engagement | Scaling up | Equity, gender, inclusion, and non-discrimination |

HH = hand hygiene

**Table S5**. Definitions of key terms.

| Implementation phase | Key terms | Definition |
|---|---|---|
| **Step 1**: prepare for action | Key leadership | Identify the key stakeholders that drive or influence hand hygiene community settings, including a hand hygiene champion (e.g., head of state, minister) |
| | Stakeholder engagement | Secure buy-in through consultation with key stakeholders |
| | Allocation of resources for planning | Identify and allocate necessary resources for implementation, including building capacity at all levels across all settings |
| | Coordination mechanisms | Create a coordination mechanism |
| | Potential barriers and challenges | Identify potential barriers and challenges to developing an action plan or programme |
| **Step 2**: analyse the situation | Undertake the situation analysis | Undertake a situation analysis of hand hygiene in community settings |
| | Data collection | Collect data on hand hygiene infrastructure and practices across community settings |
| | Data analysis | Analyse the hand hygiene data collected, disaggregated by community setting |
| | Presentation of results | Prepare and share results from baseline data collection |
| | Enabling environment | Understand governance, financing, capacity development, data and information, and innovation for hand hygiene in community settings |
| **Step 3**: develop an action plan | Stakeholder review of baseline data | Engage key stakeholders to review hand hygiene baseline data |
| | Action plan development | Develop an action plan or programme for hand hygiene in community settings |
| | Action plan support and approval | Secure support and approval for the action plan or programme by engaging key stakeholders |
| | Action plan costing | Develop a fully costed action plan or programme |
| **Step 4**: execute the action plan | Roles and responsibilities | Define roles and responsibilities for executing the action plan |
| | Action plan completion timeline | Develop a timeline for completing the action plan |

| | | |
|---|---|---|
| **Step 5**: monitor, evaluate, and course correct | Tracking progress | Track progress against national or programme targets |
| | Cyclical review and analysis | Routinely collect, analyse, and review monitoring data on what works, with whom, and what is cost effective |
| | Stakeholder engagement | Engage with key stakeholders on reviewing monitoring and evaluation results |
| | Scaling up | Strategic actions and investments to scale up hand hygiene interventions in community settings based on data collected |
| **Step 6**: cross-cutting | Equity, gender, inclusion, and non-discrimination | Address gender inequalities and the inclusion of people with disabilities in the action plan or programme |

**Table S6.** Excluded documents with reasons.

| **Author** | **Year** | **Title** | **Exclusion reason** |
|---|---|---|---|
| n/a | n.d. | Capacity needs assessment tool | 1 |
| Plan International Australia | 2020 | WASH in schools implementation guidelines | 4 |
| Ministry of Health, Ethiopia | 2017 | Baby and Mother WASH Implementation Guideline | 4 |
| Harvard Kennedy School | 2020 | Action toolkit- handwashing and other preventative measures | 6 |
| CAWST | 2017 | Community WASH Promotion- Kirkpatrick Theory of Change Evaluation Tools | 1 |
| Oxfam | 2018 | A field guide for rapid implementation of handwashing promotion in emergencies | 2 |
| Global Handwashing Partnership | 2022 | Global Handwashing Day 2022 Social Media Toolkit | 6 |
| UNICEF East Asia and Pacific Regional Office | 2021 | Hand Hygiene For All: a Resource Guide for Business | 6 |
| UNICEF et al. | 2020 | Hand Hygiene For All | 6 |
| UNICEF | 2013 | Handwashing Promotion: Monitoring and Evaluation Module | 7 |
| SNV Netherlands | 2015 | Handwashing with soap: Communication Strategy & Communication Toolkit | 6 |
| CAWST | 2014 | Technical Brief: Handwashing | 6 |

| | | | |
|---|---|---|---|
| JMP | 2021 | Working paper: Mapping and gap analysis of tools designed to collect data on hand hygiene in public spaces | 6 |
| Ministry of Health, Ethiopia | 2017 | National school water supply, sanitation, and hygiene (SWASH) implementation guideline | 4 |
| WaterAid | 2013 | Toolkit part 1: Mainstreaming equity and inclusion in WASH programmes and influencing | 1 |
| People in Need (PIN) | 2017 | Behaviour Change Toolkit for international development practitioners | 1 |
| USAID | 2020 | Water, Sanitation and Hygiene (WASH) Social and Behavior Change Communication (SBCC) | 6 |
| Ministry of Education and Sports, Uganda | 2020 | WASH in Schools (WinS) user guide | 4 |
| SDG | 2020 | Sustainable development goal country costing model for WASH | 4 |
| Sanitation and Water for All (SWA) | 2019 | Sector Ministers' Meeting: Social Media Toolkit | 6 |
| WSUP | 2014 | The Urban Programming Guide: How to design and implement an effective urban WASH programme | 6 |
| UNHCR | 2019 | UNHCR WASH Manual: Practical Guidance for Refugee Settings | 2 |
| UNICEF | 2008 | UNICEF Handwashing Module | 7 |
| UNICEF Ghana | 2015 | Water, Sanitation and Hygiene in Disaster Prone Communities in Northern Ghana: programme implementation manual | 4 |
| UN-Water | 2015 | Implementing Water, Sanitation and Hygiene (WASH): Information brief | 1 |
| UNICEF | 2017 | Choosing Public Expenditure Analytical Tools for Use in the WASH Sector | 1 |
| Safe Water Network | 2019 | WASH Toolkit | 4 |
| SHARE consortium | 2018 | WASH toolkit materials | 6 |
| World Bank | 2013 | Behavior change: overview handwashing with soap toolkit | 6 |
| USAID | 2011 | Tracking behavior and iterative planning: tools for implementers | 6 |
| USAID | 2017 | Designing for Behavior Change: A Practical Field Guide | 1 |
| Plan International | 2018 | Gender and WASH monitoring tool | 1 |
| IRC | 2017 | Costing annd budget tools | 1 |
| IRC | 2016 | Looking at WASH in non-household settings: WASH away from the home information guide | 6 |
| WaterAid | 2020 | Technical Guide for handwashing facilities in public places and buildings | 6 |
| IRC | 2014 | Mapping of water, sanitation, and hygiene sustainability tools | 3 |

| | | | |
|---|---|---|---|
| WASH Cluster State of Palestine | 2022 | WASH Assessment Toolkit | 4 |
| IRC | 2015 | WASHCost Share Guide: Analyse and share life-cycle costs | 1 |
| Water.org | 2013 | Water, Sanitation, and Microfinance toolkit 1: Introduction to Opportunities in Water, Sanitation and Hygiene Finance | 6 |
| Water.org | 2013 | Water, Sanitation, and Microfinance toolkit 2: Water, Sanitation and Hygiene Financial Product Development | 6 |
| Water.org | 2013 | Water, Sanitation, and Microfinance toolkit 3: Water, Sanitation and Hygiene Financial Product Marketing | 6 |
| Water.org | 2013 | Water, Sanitation, and Microfinance toolkit 4: WASH Finance Process Mapping, Risk Management, Pricing, Internal Audit and Controls | 6 |
| Water.org | 2013 | Water, Sanitation, and Microfinance toolkit 5: WASH Financial Product Portfolio Management | 6 |
| Live & Learn Environmental Education | n.d. | School WASH Bottleneck Analysis report | 3 |
| WaterAid & Australian Aid | 2011 | Hands Up for Hygiene: teaching hygiene behaviour in pacific schools | 4 |
| WaterAid | 2011 | How to conduct a WASH accessibility and safety audit | 4 |
| Ministry of Education and Sports, Republic of Uganda | 2020 | WinS Models that Work | 4 |
| Sustainable Development | 2015 | Advocacy toolkit: influencing the post-2015 development agenda | 1 |
| IRC International Water and Sanitation Centre | 2010 | Strengthening water, sanitation and hygiene in schools: a WASH guidance manual with a focus on South Asia | 4 |
| UNICEF | 2015 | Advancing WASH in schools monitoring: working paper | 3 |
| SWA | 2020 | WASH SDG Costing Tool | 1 |
| IRC International Water and Sanitation Centre | 2005 | The joy of learning- Participatory lesson plans on hygiene, sanitation, water, health and the environment | 6 |
| Water Supply and Sanitation Collaborative Council | 2010 | Hygiene and sanitation software | 6 |
| UNICEF Nigeria and Government of Nigeria | 2001 | Trainers Participatory Hygiene and Sanitation Promotion Manual | 7 |
| USAID / WASHplus | 2015 | Teacher’s Guide to Integrating WASH in Schools | 4 |
| USAID / WASHplus | 2015 | School WASH Facilities Operation and Maintenance Guidelines | 4 |
| UNOPS | 2016 | Technical guidance for prison planning: Technical and operational considerations based on the Nelson Mandela Rules | 6 |

| | | | |
|---|---|---|---|
| UNDP Water Governance Facility/UNICEF | 2015 | Accountability in WASH: A reference guide for programming | 1 |
| IRC / Aguaconsult | 2013 | WASH Sustainability Sector Assessment Tool | 1 |
| WaterAid | 2019 | A guide to support planning, monitoring, evaluation and learning | 1 |
| Ministry of Health, Kenya | 2022 | Implementation guidelines for rural sanitation and hygiene | 4 |
| Ministry of Health, Ethiopia | 2020 | National market-based sanitation implementation guideline | 4 |
| International Labour Organization | 2016 | WASH@Work - a self-training handbook | 7 |
| UNICEF | 2020 | Guidance on COVID-19 Back-To-Work Preparedness | 6 |
| Quality Council of India (QCI) | 2020 | Workplace Assessment for Safety and Hygiene (WASH) | 6 |

Exclusion reasons include: 1= not related to hand hygiene; 2= not related to community settings; 3= not a tool for improving hand hygiene in community settings; 4= not international; 5= published before 1990; 6= not related to at least one of the 5 implementation phases for improving hand hygiene in community settings; 7= training manual.

**Table S7**. Characteristics of included documents.

| **Author** | **Title** | **Year** | **Community setting** | **Community setting category** | **Target level** | **WASH vs hand hygiene specific** | **Objective** |
|---|---|---|---|---|---|---|---|
| Concern Worldwide | Toolkit: WASH in Schools and Learning Centres | 2021 | Schools | Institutional | Programme | WASH | Provide country teams with practical guidance on the implementation of WASH activities in schools and learning centres |
| Deutsche Gesellschaft für International Zusammenarbeit (GIZ) | Field guide: hardware for group handwashing in schools | 2013 | Schools | Institutional | Programme | Hand hygiene | Outline approaches, common mistakes, and lessons learned regarding handwashing in schools |
| EAWAG | Systematic Behavior Change in Water Sanitation and Hygiene: A practical guide using the RANAS approach | 2016 | All | All | Programme | WASH | Reference guide and cookbook to systematic behavior change in the WASH sector |

| | | | | | | | |
|---|---|---|---|---|---|---|---|
| Global Handwashing Partnership (GHP) | Clean hands for all: a toolkit for hygiene advocacy | 2018 | All | All | National | Hand hygiene | Outline tools and resources to help hygiene advocates ensure that handwashing with soap is recognized as critical to health and development |
| Global Public-Private Partnership for Handwashing (PPPHW) | Post-2015 hygiene advocacy toolkit | 2015 | All | All | National | Hand hygiene | Make the case for hygiene in the WASH sector |
| IRC International Water and Sanitation Centre | Towards effective programming for WASH in Schools | 2007 | Schools | Institutional | Programme | WASH | Describe the elements needed for scaling up programmes for water, sanitation and hygiene in schools while ensuring quality and sustainability |
| Live & Learn Environmental Education & Australian Aid | Field Guide: Three Star Approach for WASH in Schools | 2011 | Schools | Institutional | Programme | WASH | Improve the effectiveness of hygiene behaviour change programmes, while ensuring that schools meet the essential criteria |
| London School of Hygiene and Tropical Medicine (LSHTM) | Behavior centred design: a practicioner's manual | 2017 | Not specified | Not specified | Programme | WASH | Guide behaviour change programmers through the process of program development |
| Peace Corps | Water, sanitation, and hygiene in schools toolkit | 2017 | Schools | Institutional | Programme | WASH | Provide guidance, resources, and ideas to support comprehensive WASH programming that is focused on behavior change and sustainability |
| Rotary International | Water, sanitation, hygiene, education, literacy: a guide to WASH in schools | 2017 | Schools | Institutional | Programme | WASH | Help Rotary members design, deliver, and evaluate their WASH in Schools programs |
| Rotary International | Understanding the school community: tool for assessing needs and collecting data | 2017 | Schools | Institutional | Programme | WASH | Help determine the water and sanitation needs in a chosen community |
| Rotary International | WASH in schools worksheets | 2017 | Schools | Institutional | Programme | WASH | Supplement to the Guide to WASH in Schools, a resource for planning a WASH in Schools project |

| | | | | | | | |
|---|---|---|---|---|---|---|---|
| Sanitation and Water for All | Water & sanitation: how to make public investment work – a handbook for finance ministers | 2020 | Not specified | Not specified | National | WASH | Call to action for ministers of finance, with inspirational case studies and forward-looking sector perspectives |
| UN-Water | TrackFin Initiative: tracking financing to sanitation, hygiene and drinking-water at the national level – guidance document | 2017 | | | National | WASH | Outline a methodology to identify and track financing to the WASH sector to help countries track sector financing on a regular and comparable basis |
| UNICEF | Water, Sanitation and Hygiene (WASH) in Schools | 2012 | Schools | Institutional | Programme | WASH | Explore various options for effectively implementing a WASH in schools programmes |
| UNICEF | WASH Bottleneck Analysis Tool (BAT) | 2011 | All | All | National | WASH | The WASH BAT helps formulate costed and prioritized Action Plans to remove the bottlenecks that constrain the WASH sector and hinder the delivery of sustainable WASH services |
| UNICEF | Developing Water, Sanitation and Hygiene (WASH) Finance Strategies: A Guide | 2022 | All | All | National | WASH | Support governments and development partners in promoting and facilitating the development of WASH finance strategies |
| UNICEF | Handwashing Promotion: Monitoring and Evaluation Module | 2013 | All | All | Programme | Hand hygiene | Outline steps for planning and implementing monitoring and evaluation (M&E) for your handwashing promotion programme |
| UNICEF | Make it Count: Guidance on disability inclusive WASH programme data collection, monitoring and reporting | 2021 | All | All | Programme | WASH | Support the development of disability inclusive WASH data collection, monitoring, and reporting |
| UNICEF | WASH Bottleneck Analysis Tool: Country Implementation Guide | 2019 | All | All | Programme | WASH | Support people who are responsible for facilitating and organizing a workshop on the application of the WASH BAT to ensure an effective outcome |
| UNICEF | Sustainability checks: guidance to design and | 2017 | All | All | Programme | WASH | Provide brief guidance on how to carry out a Sustainability Check so |

| | | | | | | | |
|---|---|---|---|---|---|---|---|
| | implement sustainability monitoring in WASH | | | | | | that the exercise can be transitioned into national monitoring systems |
| UNICEF | Strengthening enabling environment for water, sanitation, and hygiene (WASH): guidance note | 2016 | All | All | National | WASH | Orient UNICEF country staff to support governments to strengthen the WASH enabling environment |
| UNICEF | Toolkit for Water, Sanitation and Hygiene (WASH) sector strengthening | 2016 | Not specified | Not specified | National | WASH | Provide resources and available tools for partners to get an orientation on strengthening the WASH building blocks |
| UNICEF | WASH in Schools Monitoring Package | 2011 | Schools | Institutional | National | WASH | Monitor deficit at the national level |
| UNICEF and World Health Organization | Core questions and indicators for monitoring WASH in Schools in the Sustainable Development Goals | 2016 | Schools | Institutional | National | WASH | Present recommended core questions to support harmonised monitoring of WASH in schools as part of the SDGs |
| WaterAid | Toolkit: understanding and addressing equality, non-discrimination and inclusion in water, sanitation and hygiene (WASH) work | 2017 | All | All | Programme | WASH | Support on reducing inequalities in programme and advocacy work in WASH |
| WaterAid | WASH assessment tool | 2021 | Workplace | Institutional | Programme | WASH | Provide businesses with a way to measure the current WASH status, including COVID-19 specific measures |
| WBCSD | Water, sanitation and hygiene implementation at the workplace: pledge and guiding principles | 2014 | Workplace | Institutional | Programme | WASH | Not stated |
| WHO & UNICEF | Costing tool for estimating the cost of interventions to improve hand hygiene in the domestic settings | 2021 | Household | Domestic | National | Hand hygiene | Provide governments in low- and middle-income countries with a rapid point-of-departure estimate on hand hygiene costs to inform initial stages of discussion and planning |
| WHO, UNICEF, WaterAid | Hand hygiene acceleration framework tool | 2023 | All | All | National | Hand hygiene | Track the process that a government has undergone to develop and implement a plan of action for hand |

| | | | | | | | |
|---|---|---|---|---|---|---|---|
| | | | | | | | hygiene improvement and assesses the quality of that plan |
| World Bank | The Handwashing Handbook | 2005 | All | All | Programme | Hand hygiene | Outline how handwashing behavior can be changed on a large or national scale by providing lessons from industrial marketing approaches as well as from current public health thinking |
| World Bank | Practical Guidance for Measuring Handwashing Behavior | 2010 | All | All | Programme | Hand hygiene | Describe the positive and negative attributes of various commonly applied and novel methods of measuring handwashing behavior |
| World Bank, UNICEF | Toolkit on hygiene, sanitation, and water in schools | 2005 | Schools | Institutional | Programme | WASH | Help sector professionals to develop solutions that are most appropriate for the sector needs |
| World Vision | Water, Sanitation and Hygiene (WASH) Project Model | 2022 | Schools, households, and communities | All | Programme | WASH | Provide a set of evidence-based practices for the sustained provision of safe water, dignified sanitation, and good hygiene practice across a variety of contexts. |
| World Vision | Behavior change: Practical implementation guidance for programs | 2021 | Not specified | Not specified | Programme | WASH | Strengthen the process of designing behavior-change content to facilitate change for essential WASH behaviors |

**Table S8**. Implementation guidance and tools mapped to the six implementation steps and their parameters.

| Author | Title | Year | Step 1 | | | | | Step 2 | | | | | Step 3 | | | | | Step 4 | | Step 5 | | | | Step 6 |
|---|---|---|---|---|---|---|---|---|---|---|---|---|---|---|---|---|---|---|---|---|---|---|---|---|
| | | | a | b | c | d | e | a | b | c | d | e | a | b | c | d | e | a | b | a | b | c | d | e |
| World Bank | The Handwashing Handbook | 2005 | | | | | | | | | | | | | | | | | | | | | | |
| World Bank, UNICEF | Toolkit on hygiene, sanitation, and water in schools | 2005 | | | | | | | | | | | | | | | | | | | | | | |
| GHP | Clean hands for all: a toolkit for hygiene advocacy | 2018 | | | | | | | | | | | | | | | | | | | | | | |
| UNICEF | Water, Sanitation and Hygiene (WASH) in Schools | 2012 | | | | | | | | | | | | | | | | | | | | | | |
| Peace Corps | Water, sanitation, and hygiene in schools toolkit | 2017 | | | | | | | | | | | | | | | | | | | | | | |
| WaterAid | Toolkit: understanding and addressing equality, non-discrimination and inclusion in water, sanitation and hygiene (WASH) work | 2017 | | | | | | | | | | | | | | | | | | | | | | |
| WaterAid | WASH assessment tool | 2021 | | | | | | | | | | | | | | | | | | | | | | |
| WHO, UNICEF, WaterAid | Hand hygiene acceleration framework tool | 2023 | | | | | | | | | | | | | | | | | | | | | | |
| PPPHW | Post-2015 hygiene advocacy toolkit | 2015 | | | | | | | | | | | | | | | | | | | | | | |
| World Bank | Practical Guidance for Measuring Handwashing Behavior | 2010 | | | | | | | | | | | | | | | | | | | | | | |
| UNICEF | Developing Water, Sanitation and Hygiene (WASH) Finance Strategies: A Guide | 2022 | | | | | | | | | | | | | | | | | | | | | | |
| UNICEF | Handwashing Promotion: Monitoring and Evaluation Module | 2013 | | | | | | | | | | | | | | | | | | | | | | |

| | | |
|---|---|---|
| UNICEF | Make it Count: Guidance on disability inclusive WASH programme data collection, monitoring and reporting | 2021 |
| Concern Worldwide | Toolkit: WASH in Schools and Learning Centres | 2021 |
| UNICEF | WASH Bottleneck Analysis Tool: Country Implementation Guide | 2019 |
| UNICEF | Sustainability checks: guidance to design and implement sustainability monitoring in WASH | 2017 |
| UNICEF | Strengthening enabling environment for water, sanitation, and hygiene (WASH): guidance note | 2016 |
| World Vision | Water, Sanitation and Hygiene (WASH) Project Model | 2022 |
| WHO | Costing tool for estimating the cost of interventions to improve hand hygiene in the domestic settings | 2021 |
| World Vision | Behavior change: Practical implementation guidance for programs | 2021 |
| EAWAG | Systematic Behavior Change in Water Sanitation and Hygiene: A practical guide using the RANAS approach | 2016 |
| LSHTM | Behavior centred design: a practicioner's manual | 2017 |

| | | |
|---|---|---|
| UNICEF | Toolkit for Water, Sanitation and Hygiene (WASH) sector strengthening | 2016 |
| Live & Learn Environmental Education & Australian Aid | Field Guide: Three Star Approach for WASH in Schools | 2011 |
| UNICEF | WASH in Schools Monitoring Package | 2011 |
| UN-Water | TrackFin Initiative: tracking financing to sanitation, hygiene and drinking-water at the national level - guidance document | 2017 |
| GIZ | Field guide: hardware for group handwashing in schools | 2013 |
| IRC International Water and Sanitation Centre | Towards effective programming for WASH in Schools | 2007 |
| WBCSD | Water, sanitation and hygiene implementation at the workplace - Pledge and guiding principles | 2014 |
| Sanitation and Water for All | Water & sanitation: how to make public investment work - a handbook for finance ministers | 2020 |
| UNICEF and World Health Organization | Core questions and indicators for monitoring WASH in Schools in the SDGs | 2016 |
| Rotary International | Water, sanitation, hygiene, education, literacy: a guide to WASH in schools | 2017a |

| | | |
|---|---|---|
| Rotary International | Understanding the school community: tool for assessing needs and collecting data | 2017b |
| Rotary International | WASH in schools worksheets | 2017c |
| UNICEF | WASH Bottleneck Analysis Tool (BAT) | 2011 |

Light grey shading indicates implementation guidance and dark grey shading indicates a tool.
